# Association of daily physical activity, handgrip strength and visceral adiposity with mortality, cardiovascular events and cancers in Japanese individuals with diabetes

**DOI:** 10.64898/2026.06.09.26355239

**Authors:** Hidetaka Hamasaki, Vicky Taxiarchi

**Author notes:** Corresponding author: Hidetaka Hamasaki, Japanese Academy of Health and Practice, The 3rd floor of Mutsumi Building, 2-10-48, Kitasaiwai, Nishi-ku, Yokohama, Kanagawa 220-0004, Japan, Hamasaki Clinic, Ebina, Kanagawa, Japan, Email address (H. Hamasaki).

## Abstract

**Aims:** This study examined the associations of body composition and daily physical activity with mortality, CV events and cancer in individuals with diabetes.

**Methods:** This prospective cohort study included individuals with diabetes treated at a specialised clinic in Japan between January 2018 and March 2023. Body composition, including visceral adipose tissue (VAT), was assessed by bioelectrical impedance analysis. Daily physical activity was evaluated using the non-exercise activity thermogenesis (NEAT) questionnaire, and handgrip strength (HGS) was measured by dynamometry. Cox proportional hazards models were used to assess associations with mortality, CV events, and cancer.

**Results:** Among 2,024 participants (mean age 63.0 years, BMI 24.6 kg/m², HbA1c 7.8%), NEAT score, HGS and VAT were not independently associated with all-cause mortality. Higher VAT was associated with increased cancer risk (HR = 1.288; 95% CI, 1.008–1.646; p = 0.043). Higher HGS was inversely associated with CV event risk (HR = 0.955; 95% CI, 0.925–0.986; p = 0.005). NEAT score was not associated with any outcome.

**Conclusions:** Higher VAT was associated with increased cancer risk, whereas higher HGS was associated with a lower rate of CV events. Incorporating body composition and HGS assessments into clinical practice may provide additional prognostic information in individuals with diabetes.

## 1. Introduction

Ageing is associated with various physiological changes, including decreased mobility, impaired balance and reduced lower extremity strength, which elevate the risk of falls [1]. These changes increase susceptibility to non-communicable diseases, including diabetes, cardiovascular (CV) disease and cancer [2], partly through declining lean body mass and rising visceral adipose tissue, which impair pancreatic β-cell function and promote T2D via inflammation and oxidative stress. Consequently, the prevalence and burden of T2D are expected to increase [3,4]. Obesity is also increasing across all age groups, sexes, and populations, with the highest prevalence observed among older adults [5].

Recently, skeletal muscle has been recognised as an endocrine organ that secretes myokines influencing glucose and lipid metabolism, cognitive function, bone formation and vascular health [6]. Sarcopenic obesity, defined by the concurrent presence of muscle loss and fat accumulation, has emerged as a critical health concern and is associated with increased mortality [7]. Age-related changes in body composition, together with rising obesity rates, contribute to its growing prevalence [8]. A meta-analysis reported that sarcopenic obesity increases the risk of T2D by approximately 38% compared with overweight or obesity alone [9]. However, commonly used anthropometric indicators such as body weight or body mass index (BMI) cannot accurately differentiate muscle, fat and bone mass, and therefore fail to identify sarcopenia or sarcopenic obesity. Although the so-called obesity paradox suggests that moderate overweight may be associated with better prognoses for CV disease, cancer and overall survival in older adults [10–12], it is well established that excess adipose tissue exerts adverse effects on health outcomes [13,14]. Therefore, objective assessments of body composition, including body fat, skeletal muscle mass (SMM) and visceral adiposity, are essential for optimising treatment strategies and improving outcomes in individuals with diabetes, sarcopenia and sarcopenic obesity.

Increasing physical activity (PA) plays an important role in mitigating sarcopenic obesity in individuals with diabetes [15]. Walking is the most representative form of daily PA, and a meta-analysis of 24 randomized controlled trials demonstrated that walking interventions reduced body weight and body fat [16]. PA was also associated with lower mortality and fewer major CV events [17]. Highly active individuals had reduced risks of diabetes, CV diseases and several types of cancer [18]. In addition to structured exercise, daily PA significantly improves health outcomes. However, few studies have separately evaluated daily PA and regular exercise, and investigated their associations with CV events and cancer incidence in individuals with diabetes.

However, few studies have simultaneously examined daily non-exercise PA, muscle strength, and visceral adiposity in relation to major clinical outcomes among individuals with diabetes. This study therefore aimed to examine the associations of NEAT score, HGS and body composition measures with all-cause mortality, CV events and incident cancer.

## 2. Methods

### 2.1 Study design and subjects

We conducted a prospective observational study involving individuals with diabetes treated at Hamasaki Clinic. Between January 2018 and March 2023, all individuals with diabetes attending the clinic for routine examinations were invited to participate. Those who provided consent were consecutively enrolled, resulting in a study population of 2,024 participants. Participants were followed from their first visit until death or 31 March 2023. The study protocol received approval from the Japan Medical Association Ethical Review Board (reference number: JMA-IIA00340) and adhered to the principles of the Declaration of Helsinki. Information regarding the study procedures, personal information protection and verbal consent from participants were documented in the medical records. Moreover, participants were provided with an opportunity to opt out of the study.

The following were the exclusion criteria: participants aged < 20 years, those with internal metallic implants (e.g., pacemakers), those who underwent upper or lower limb amputation surgery and those who could not maintain a standing position owing to a decline in activities of daily living because their body composition could not be accurately measured using the bioelectrical impedance analysis (BIA) device (seca mBCA 515; seca GmbH & Co. KG., Hamburg, Germany). Participants with a history of cancer at baseline were also excluded, ensuring that only cancers newly diagnosed during follow-up were classified as incident events. Participants with a history of CV disease were not excluded; instead, baseline CV disease was included as a covariate in the multivariable models.

### 2.2 Data collection method

The baseline data were collected at the first visit to the clinic during the study period. Participants’ characteristics including age, sex, height, weight, BMI, waist circumference (WC), treatment duration, blood pressure, HGS, blood data, disease conditions including diabetic neuropathy and nephropathy markers, Non-exercise activity thermogenesis (NEAT) score and body composition data were investigated through medical record reviews. Data collection occurred on consultation days as part of routine medical practice at the clinic.

### 2.3 Body composition analysis

Using the seca medical Body Composition Analyser 515; seca mBCA 515 (seca GmbH & Co. KG., Hamburg, Germany), body composition was assessed. The seca mBCA 515 uses an eight-electrode system (four pairs on hands and feet) to measure segmental impedance in a standing position. Resistance (R) and reactance (Xc) were measured at 50 kHz, and the phase angle (φ) was derived accordingly. Participants avoided exercise (12 h) and alcohol (24 h) pre-measurement [19]. Manufacturer accuracy at 5 and 50 kHz is ±5Ω (impedance) and 0.5° (phase angle) (seca GmbH & Co. KG., 2016). SMM, fat-free mass (FFM), fat mass (FM), total body water and extracellular water of the whole body, trunk and extremities were calculated on the basis of the body impedance analysis. The SMM index (SMI) was calculated as the ratio of SMM in kilograms to the square of body height in meters. Moreover, visceral adipose tissue volume (VAT) was estimated using the segmental impedance measurements. VAT estimated using this BIA device was highly correlated with VAT (r² = 0.8) measured using MRI [20].

### 2.4 Daily physical activity

Daily PA was evaluated using a validated questionnaire assessing NEAT in individuals in Japan [21]. Using a questionnaire adapted from a PA compilation, healthcare providers in the clinic asked participants about their NEAT-related PA routines. The NEAT score was developed by consulting an article from The Compendium of Physical Activities [22]. The questionnaire was designed to assess participants’ usual habitual activities, excluding gym workouts and other regular exercise routines. The questionnaire comprised the following two parts: locomotive activities (11 items) and non-locomotive activities (25 items). Each questionnaire item was assessed on a scale of 1–3 points on the basis of the degree of daily PA, and the scores were subsequently aggregated to determine the NEAT score (maximum, 108 points) (Supplementary file).

### 2.5 Handgrip strength

HGS was measured using a Smedley hand dynamometer (MIS, Tokyo, Japan) with participants in a standing position. Two measurements were obtained for each hand, and the mean of all four measurements was used for analysis (kg).

### 2.6 Medical history taking

During the first medical examination of the study period, participants were asked about their past medical history, diabetes duration and social history, including regular exercise, smoking and drinking habits. Diabetes duration was calculated from the diagnosis date to the data collection date (the first medical examination during the study period), and the results were converted into years. Smoking and drinking habits were recorded as binary variables (yes/no).

### 2.7 Diet

During the initial visit, participants received guidance from a certified nutritional educator to follow a calorie-restricted diet of 25–30 kcal/kg (based on the ideal body weight), with approximately 50%–60% and 20%–30% of energy from carbohydrates and fat, respectively, and 1.0–1.2 g/kg of protein per day, as a dietary therapy for diabetes [23]. This prescribed diet therapy was to be maintained throughout the study period. Adherence was evaluated during each clinic visit, and participants were instructed to modify their dietary intake when the Japanese clinical practice guideline for diabetes was not followed [23].

### 2.8 Anthropometric and physiological measurements

Body height was measured using a rigid stadiometer (seca 217; seca Nihon Co., Ltd., Chiba, Japan), and body weight was determined using calibrated scales (seca 899; seca Nihon Co., Ltd., Chiba, Japan). BMI was calculated as the ratio of body weight in kilograms to the square of body height in meters. WC was measured at the umbilical level during the exhalation phase with participants in a standing position. Blood pressure was measured with a sphygmomanometer while participants were seated.

### 2.9 Biochemical measurements

The study also included measurements of biochemical markers, including serum total cholesterol, triglycerides, high-density lipoprotein cholesterol, low-density lipoprotein cholesterol, plasma glucose and haemoglobin A1c (HbA1c) levels. Serum C-peptide levels were also measured using a chemiluminescent immunoassay kit.

### 2.10 Outcomes

The occurrence of deaths, CV events (heart failure, stroke, angina pectoris and myocardial infarction) and cancers was confirmed through reports from other medical institutions and interviews with participants or their families during regular clinic visits. The CV outcome was defined as the first CV event recorded after baseline, irrespective of any history of CV disease. Alternatively, information was gathered through patient referral documents from hospitals where participants had been admitted when they died or developed severe diseases.

### 2.11 Statistical analysis

Statistical analyses were performed using R version 4.6.0 (R Foundation for Statistical Computing, Vienna, Austria). Continuous values were expressed as means ± standard deviations (SDs), whereas categorical variables were presented as numbers or percentages.

The cross-sectional associations of NEAT score, HGS, and VAT with anthropometric, physiological, biochemical and body composition parameters were examined using univariable and multivariable linear regression.

Multivariable restricted cubic spline models were used to assess potential non-linear associations between each exposure and the outcomes. As no significant evidence of non-linearity was observed, the associations of NEAT score, HGS and VAT with all-cause mortality, CV events and incident cancer were subsequently evaluated using Cox proportional hazards models. Results are presented as hazard ratios (HRs) with 95% confidence intervals (CIs).

Separate models were fitted for each exposure. Follow-up began on the date of exposure assessment and continued until the first occurrence of the outcome of interest, death or 31 March 2023. Analyses were restricted to participants with complete data on the relevant exposure, outcome and covariates. Three progressively adjusted models were specified for CV events and cancer incidence. Model 1 included age and sex; the NEAT and HGS models were additionally adjusted for body mass index. The VAT models were not adjusted for BMI because the BIA-derived VAT estimate incorporates body weight and height [20], which are also used to calculate BMI; a BMI-adjusted VAT model was therefore examined as a sensitivity analysis. Model 2 further included smoking status, diabetes duration, and a history of CV disease, and Model 3 additionally included regular exercise. Because of the limited number of deaths, analyses of all-cause mortality were adjusted for age and sex only.

Each exposure was modelled as a continuous variable, and HRs were estimated per 1-point increase in NEAT score, per 1-kg increase in HGS and per 1-L increase in VAT. For CV events and cancer incidence, Fine-Gray subdistribution hazard models were also fitted, with death from any cause treated as a competing event. The proportional hazards assumption was evaluated using tests based on Schoenfeld residuals. In a sensitivity analysis, participants with a history of CV disease at baseline were excluded to restrict the outcome to first-ever CV events, and the interactions between each exposure and baseline CV disease history were tested. All tests were two-sided, and P values < 0.05 were considered statistically significant.

## 3. Results

### 3.1 Participants’ characteristics at baseline

This study included 2,024 individuals (1,176 men and 848 women) with diabetes. The mean age of the study participants was 63.0 ± 13.2 years, with a mean BMI of 24.6 ± 4.4 kg/m². The mean plasma glucose and HbA1c values were 165.2 ± 73.5 mg/dL and 7.8% ± 1.9%, respectively, indicating relatively poor glycaemic control. Among the study participants, 1,504, 1,472 and 1,106 had measurements for NEAT score, HGS and body composition, respectively. Notably, 385 participants (19%) had a CV disease history. The mean values for NEAT scores, HGS and VAT were 62.6 ± 10.8 points, 27.1 ± 10.3 kg and 3.1 ± 1.2 L, respectively. Demographic and clinical data at baseline are detailed in Table 1.

**Table 1.**
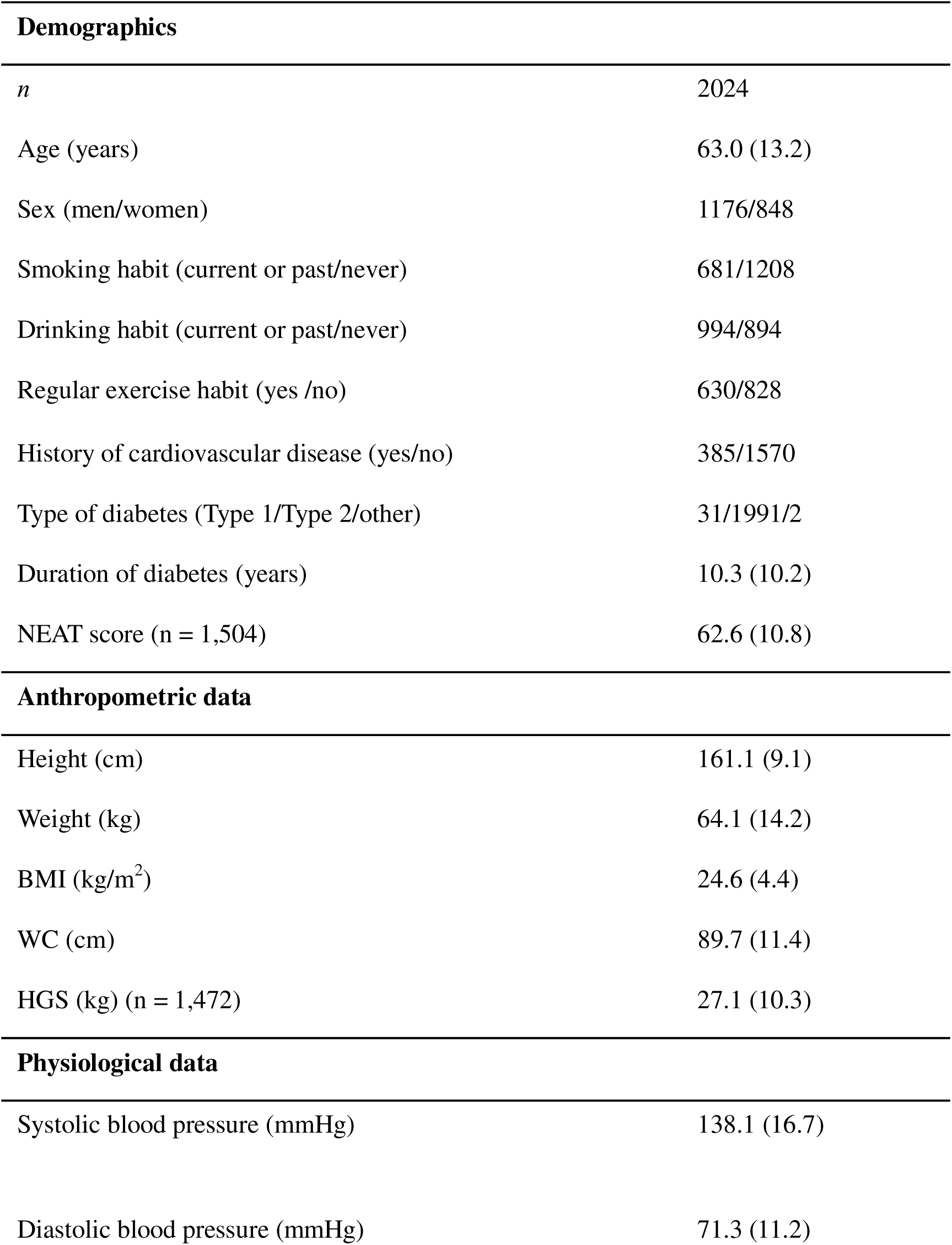
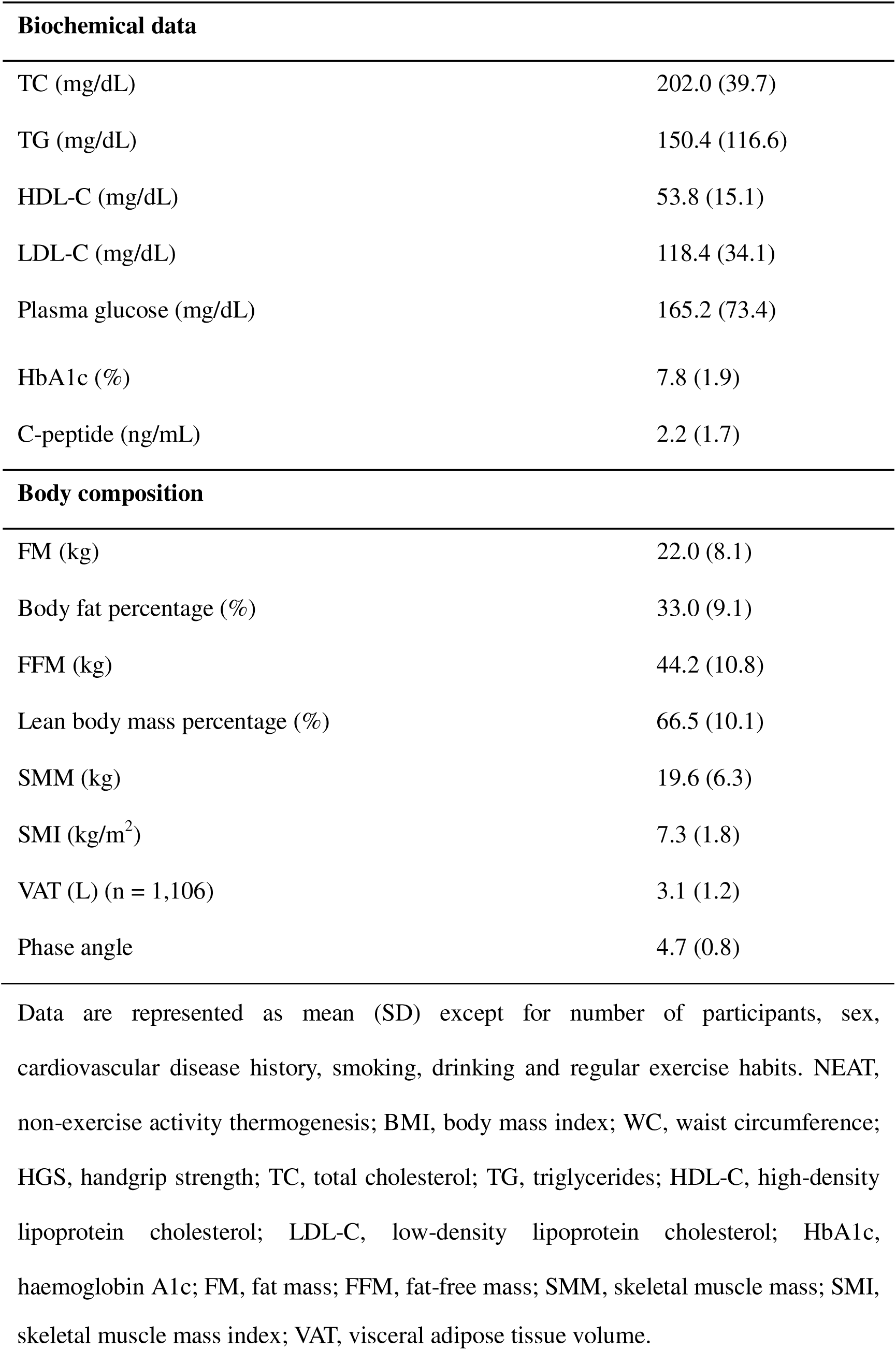
Characteristics of participants.

| **Demographics** |  |
| --- | --- |
| *n* | 2024 |
| Age (years) | 63.0 (13.2) |
| Sex (men/women) | 1176/848 |
| Smoking habit (current or past/never) | 681/1208 |
| Drinking habit (current or past/never) | 994/894 |
| Regular exercise habit (yes /no) | 630/828 |
| History of cardiovascular disease (yes/no) | 385/1570 |
| Type of diabetes (Type 1/Type 2/other) | 31/1991/2 |
| Duration of diabetes (years) | 10.3 (10.2) |
| NEAT score (n = 1,504) | 62.6 (10.8) |
| **Anthropometric data** |  |
| Height (cm) | 161.1 (9.1) |
| Weight (kg) | 64.1 (14.2) |
| BMI (kg/m<sup>2</sup>) | 24.6 (4.4) |
| WC (cm) | 89.7 (11.4) |
| HGS (kg) (n = 1,472) | 27.1 (10.3) |
| **Physiological data** |  |
| Systolic blood pressure (mmHg) | 138.1 (16.7) |
| Diastolic blood pressure (mmHg) | 71.3 (11.2) |

| **Biochemical data** |  |
| --- | --- |
| TC (mg/dL) | 202.0 (39.7) |
| TG (mg/dL) | 150.4 (116.6) |
| HDL-C (mg/dL) | 53.8 (15.1) |
| LDL-C (mg/dL) | 118.4 (34.1) |
| Plasma glucose (mg/dL) | 165.2 (73.4) |
| HbA1c (%) | 7.8 (1.9) |
| C-peptide (ng/mL) | 2.2 (1.7) |
| **Body composition** |  |
| FM (kg) | 22.0 (8.1) |
| Body fat percentage (%) | 33.0 (9.1) |
| FFM (kg) | 44.2 (10.8) |
| Lean body mass percentage (%) | 66.5 (10.1) |
| SMM (kg) | 19.6 (6.3) |
| SMI (kg/m<sup>2</sup>) | 7.3 (1.8) |
| VAT (L) (n = 1,106) | 3.1 (1.2) |
| Phase angle | 4.7 (0.8) |

Data are represented as mean (SD) except for number of participants, sex, cardiovascular disease history, smoking, drinking and regular exercise habits. NEAT, non-exercise activity thermogenesis; BMI, body mass index; WC, waist circumference; HGS, handgrip strength; TC, total cholesterol; TG, triglycerides; HDL-C, high-density lipoprotein cholesterol; LDL-C, low-density lipoprotein cholesterol; HbA1c, haemoglobin A1c; FM, fat mass; FFM, fat-free mass; SMM, skeletal muscle mass; SMI,
skeletal muscle mass index; VAT, visceral adipose tissue volume.

### 3.2 Associations between daily physical activity, handgrip strength and body composition

Multiple regression analyses demonstrated that NEAT score was positively associated with HGS (β = 0.06, p = 0.005) and lean body mass percentage (β = 0.09, p = 0.004), whereas it was negatively associated with FM (β = −0.06, p = 0.003) and body fat percentage (β = −0.06, p = 0.014) after adjusting for covariates. However, no significant associations were observed between the NEAT score and SMM, FFM, VAT and phase angle (Table 2).

**Table 2.**
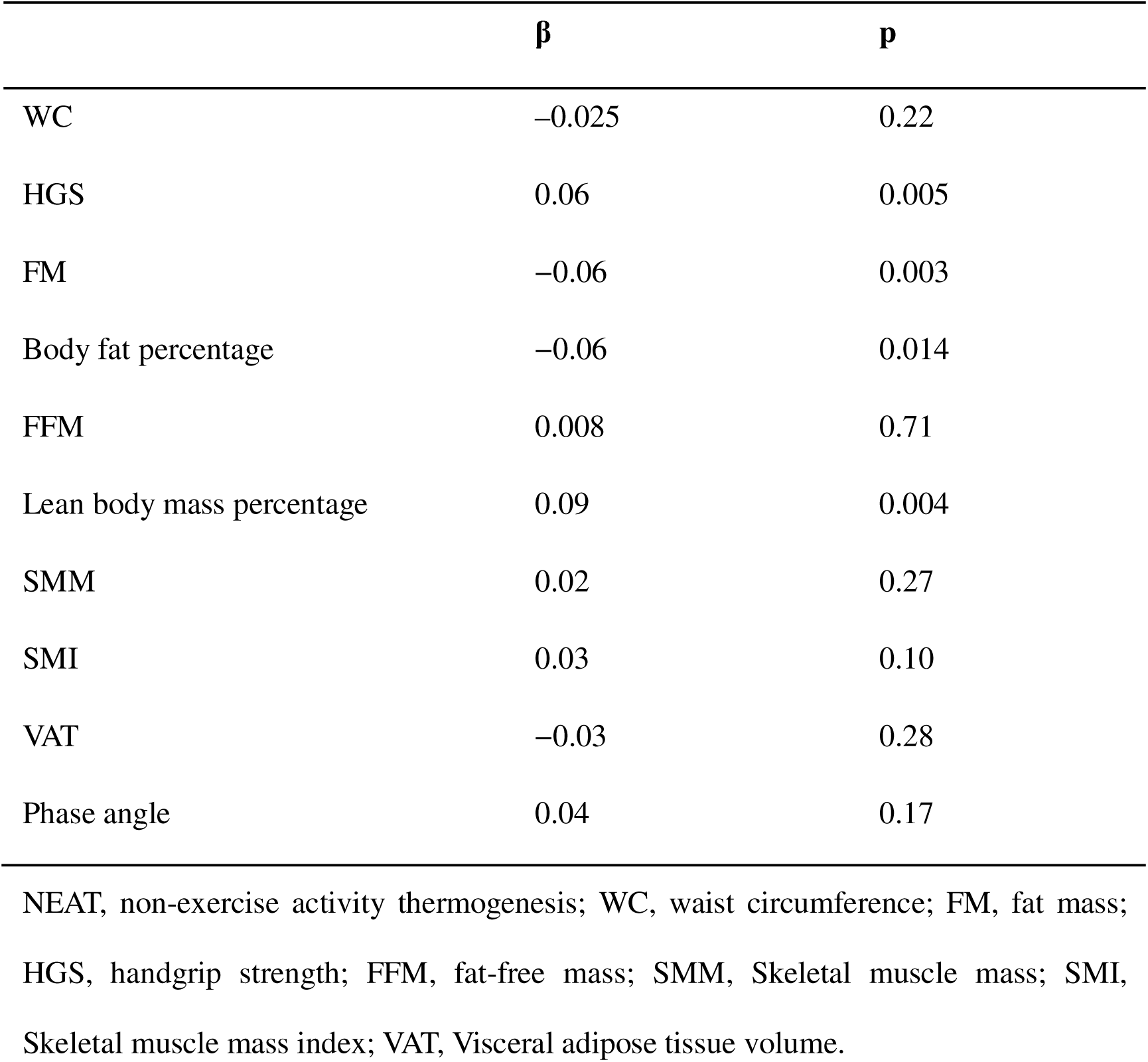
Associations of NEAT score with HGS and body composition.

### 3.3 All-cause mortality

During a mean follow-up of 4.09 ± 1.61 years, 44 (2.2%) participants died, 204 (10.1%) experienced CV events and 155 (7.7%) developed cancer.

Univariate Cox proportional hazards analyses showed that NEAT score (HR = 0.987; 95% CI, 0.956–1.019; p = 0.41) and VAT (HR = 0.547; 95% CI, 0.259–1.156; p = 0.11) were not associated with all-cause mortality, whereas higher HGS was associated with reduced all-cause mortality risk (HR = 0.938; 95% CI, 0.900–0.978; p = 0.003). After adjustment for age and sex, NEAT score, HGS and VAT were not associated with all-cause mortality (Table 3). For VAT, the age- and sex-adjusted analysis was based on only 10 deaths, and VAT was not associated with all-cause mortality (HR = 0.577; 95% CI, 0.235–1.421; p = 0.23). In separate age- and sex-adjusted models, higher SMM (HR = 0.803; 95% CI, 0.654–0.985; p = 0.035), SMI (HR = 0.449; 95% CI, 0.266–0.760; p = 0.003) and phase angle (HR = 0.171; 95% CI, 0.066–0.441; p < 0.001) were associated with lower all-cause mortality, whereas FM and FFM were not (Supplementary Table 2).

**Table 3.** Cox proportional hazards analysis for evaluating the associations of NEAT score, HGS and VAT with all-cause mortality.

|  | NEAT score |  |  | HGS |  |  | VAT |  |  |
| --- | --- | --- | --- | --- | --- | --- | --- | --- | --- |
|  | HR | 95% CI | p | HR | 95% CI | p | HR | 95% CI | p |
| Crude model | 0.987 | 0.956–<br>1.019 | 0.41 | 0.938 | 0.900–<br>0.978 | 0.003 | 0.547 | 0.259–<br>1.156 | 0.11 |
| Adjusted for age and sex | 0.984 | 0.954–<br>1.015 | 0.32 | 0.981 | 0.920–<br>1.045 | 0.55 | 0.577 | 0.235–<br>1.421 | 0.23 |

NEAT, non-exercise activity thermogenesis; HGS, handgrip strength; VAT, visceral adipose tissue volume; HR, hazard ratio; CI, confidence interval. Hazard ratios are expressed per 1-point increase in NEAT score, per 1-kg increase in HGS, and per 1-L increase in VAT. The number of participants (deaths) was 1,463 (32) for NEAT, 1,414 (28) for HGS, and 1,103 (10) for VAT.

### 3.4 Cardiovascular events

Univariate Cox proportional hazards analyses showed that NEAT score (HR = 0.998; 95% CI, 0.980–1.016; p = 0.81) and VAT (HR = 1.143; 95% CI, 0.910–1.435; p = 0.25) were not associated with CV events, whereas participants with higher HGS were less likely to experience CV events (HR = 0.961; 95% CI, 0.941–0.982; p < 0.001). Following multivariable adjustment, higher HGS remained significantly associated with reduced CV event risk (HR = 0.955; 95% CI, 0.925–0.986; p = 0.005) (Table 4). In Fine-Gray competing risks analysis, this association remained significant (subdistribution HR = 0.955; 95% CI, 0.926–0.985; p = 0.004). In a sensitivity analysis restricted to participants without a history of CV disease at baseline, thereby limiting the outcome to first-ever CV events, the association between HGS and CV events was no longer statistically significant (60 events among 950 participants; HR = 0.967; 95% CI, 0.931–1.006; p = 0.096). The interaction between HGS and baseline CV disease history was not significant (p = 0.41), providing no evidence that the association differed according to the presence or absence of pre-existing CV disease. When the NEAT and VAT analyses were similarly restricted to first-ever CV events, the conclusions remained unchanged. None of the other body composition indices (FM, FFM, SMM, SMI or phase angle) was significantly associated with CV events in either model (Supplementary Table 2).

**Table 4.** Cox proportional hazards analysis for evaluating the associations of NEAT score, HGS and VAT with cardiovascular events.

|  | NEAT score |  |  | HGS |  |  | VAT |  |  |
| --- | --- | --- | --- | --- | --- | --- | --- | --- | --- |
|  | HR | 95% CI | p | HR | 95% CI | p | HR | 95% CI | p |
| Crude model | 0.998 | 0.980–1.016 | 0.81 | 0.961 | 0.941–<br>0.982 | <0.001 | 1.143 | 0.910–<br>1.435 | 0.25 |
| Model 1 | 1.000 | 0.981–1.019 | 0.99 | 0.947 | 0.917–<br>0.977 | 0.001 | 1.186 | 0.928–<br>1.516 | 0.17 |
| Model 2 | 1.008 | 0.989–1.027 | 0.40 | 0.955 | 0.925–<br>0.986 | 0.005 | 1.175 | 0.903–<br>1.527 | 0.23 |
| Model 3 | 1.010 | 0.991–1.029 | 0.32 | 0.956 | 0.924–<br>0.988 | 0.008 | 1.038 | 0.763–<br>1.414 | 0.81 |

NEAT, non-exercise activity thermogenesis; HGS, handgrip strength; VAT, visceral adipose tissue volume; HR, hazard ratio; CI, confidence interval. Model 1 was adjusted for age, sex, and BMI. Model 2 was additionally adjusted for smoking, diabetes duration, and a history of CV disease. Model 3 was additionally adjusted for regular exercise. The VAT models were not adjusted for BMI. Hazard ratios are expressed per 1-point increase in NEAT score, per 1-kg increase in HGS, and per 1-L increase in VAT. The numbers of participants (events) in the crude, Model 1, Model 2, and Model 3 analyses were, respectively, 1,281 (104), 1,266 (104), 1,162 (104), and 1,145 (103) for NEAT; 1,237 (89), 1,237 (89), 1,171 (88), and 998 (78) for HGS; and 974 (46), 973 (46), 852 (45), and 696 (40) for VAT.

### 3.5 Cancer

Univariate Cox proportional hazards analyses showed that NEAT score (HR = 1.001; 95% CI, 0.981–1.021; p = 0.96) and VAT (HR = 1.169; 95% CI, 0.926–1.475; p = 0.19) were not associated with cancer incidence, whereas HGS was associated with decreased cancer incidence (HR = 0.973; 95% CI, 0.950–0.997; p = 0.025). Following multivariable adjustment, higher VAT was significantly associated with increased cancer risk (HR = 1.288; 95% CI, 1.008–1.646; p = 0.043) (Table 5). In Fine-Gray competing risks analysis, this association was confirmed (subdistribution HR = 1.288; 95% CI, 1.047–1.584; p = 0.016). NEAT score and HGS were not significantly associated with cancer risk. Apart from VAT, none of the other body composition indices was significantly associated with cancer incidence in either model (Supplementary Table 2).

**Table 5.** Cox proportional hazards analysis for evaluating the associations of NEAT score, HGS and VAT with the development of cancer.

|  | NEAT score |  |  | HGS |  |  | VAT |  |  |
| --- | --- | --- | --- | --- | --- | --- | --- | --- | --- |
|  | HR | 95% CI | p | HR | 95% CI | p | HR | 95% CI | p |
| Crude model | 1.001 | 0.981–1.021 | 0.96 | 0.973 | 0.950–0.997 | 0.025 | 1.169 | 0.926–1.475 | 0.19 |
| Model 1 | 1.007 | 0.987–1.028 | 0.48 | 0.995 | 0.958–1.034 | 0.80 | 1.295 | 1.015–1.652 | 0.038 |
| Model 2 | 1.010 | 0.989–1.031 | 0.36 | 1.000 | 0.963–1.040 | 0.99 | 1.288 | 1.008–1.646 | 0.043 |
| Model 3 | 1.006 | 0.985–1.028 | 0.57 | 0.987 | 0.946–1.029 | 0.53 | 1.345 | 1.067–1.695 | 0.012 |

NEAT, non-exercise activity thermogenesis; HGS, handgrip strength; VAT, visceral adipose tissue volume; HR, hazard ratio; CI, confidence interval. Model 1 was adjusted for age, sex, and BMI. Model 2 was additionally adjusted for smoking, diabetes duration, and a history of CV disease. Model 3 was additionally adjusted for regular exercise. The VAT models were not adjusted for BMI. Hazard ratios are expressed per 1-point increase in NEAT score, per 1-kg increase in HGS, and per 1-L increase in VAT. The numbers of participants (events) in the crude, Model 1, Model 2, and Model 3 analyses were, respectively, 1,414 (82), 1,398 (82), 1,291 (80), and 1,275 (80) for NEAT; 1,355 (69), 1,354 (69), 1,280 (68), and 1,095 (58) for HGS; and 1,066 (39), 1,065 (39), 938 (39), and 771 (34) for VAT.

## 4. Discussion

This study demonstrated that higher VAT was associated with an increased risk of cancer, whereas greater HGS was associated with a lower risk of CV events. VAT was not significantly associated with all-cause mortality, and NEAT score was not significantly associated with any of the three outcomes.

Current evidence links excess body fat to an increased risk of multiple cancer types. Although the precise mechanisms remain unclear, insulin resistance, adipokine dysregulation, oxidative stress and chronic inflammation contribute to cancer progression [24]. Iyengar et al. [25] found that the highest trunk fat mass quartile increased invasive breast cancer risk in postmenopausal women without obesity. Katzmarzyk et al. [26] reported associations between VAT and total FM with histologically confirmed invasive cancer. A Mendelian randomisation study by Xu et al. found that genetically predicted VAT and liver fat, but not BMI, were associated with an increased risk of primary liver cancer [27]. An umbrella review of observational and Mendelian randomisation studies found robust observational evidence linking T2D to colorectal, hepatocellular, gallbladder, breast, endometrial and pancreatic cancers. The review also identified potential causal associations of genetically predicted T2D or fasting insulin concentrations with several of these cancers [28]. In this study, higher VAT was associated with higher serum C-peptide levels, suggesting greater insulin resistance among individuals with diabetes and high VAT, which may influence cancer incidence (Supplementary Table 3). In our data, higher VAT was consistently associated with a higher hazard of cancer across all adjusted models, and the association remained evident in the competing-risk analysis. It was no longer statistically significant only after additional adjustment for BMI (HR = 1.227; 95% CI, 0.865–1.739; p = 0.25), although the point estimate remained above unity. The VAT value used in this study was not measured directly but was estimated by bioelectrical impedance analysis using anthropometric and impedance parameters, including body weight and height, which are also used to calculate BMI. Adjustment for BMI therefore partly conditions on information already incorporated into the exposure estimate. This may reduce precision and change the estimand, as the BMI-adjusted coefficient reflects the association of VAT relative to a given body size rather than the overall association between VAT and cancer. Because body weight, height, and VAT were assessed concurrently, and because BMI reflects a combination of adiposity, muscle mass, and body frame, we did not consider BMI to be a straightforward confounder. We therefore treated the model without BMI adjustment as the primary analysis and the BMI-adjusted model as a sensitivity analysis. BMI itself was not associated with cancer incidence (HR = 1.03; p = 0.60), and the variance inflation factor did not indicate severe multicollinearity. Nevertheless, because the VAT estimation algorithm is proprietary and the two models address different research questions, the observed association should be interpreted with caution.

Diabetes and low HGS are associated with increased risks of mortality and CV events [29]. The UK Biobank cohort study found that individuals with diabetes and low HGS had a higher CV mortality risk than those with high HGS [30]. Similarly, in this study, each 1-kg increase in HGS was associated with a 5% lower CV event risk. The role of HGS in reducing CV risk remains unclear; however, a two-sample Mendelian randomisation study found that genetic predisposition to higher HGS had a favourable effect on coronary heart disease and myocardial infarction [31]. Myokines such as IL-6, irisin and follistatin-like protein 1 may promote myocardial tissue regeneration and counteract age-related muscle decline [32]. Endocrine hormones, including testosterone and ghrelin, together with myokines, may exert CV-protective effects by modulating autophagy, apoptosis, insulin resistance and inflammation [33]. Although previous studies have associated higher HGS with a more favourable prognosis [29,34], we found no significant association between HGS and all-cause mortality or cancer incidence. The relatively short follow-up duration (mean 4.09 years) may explain this. A meta-analysis of 48 studies examining HGS and mortality reported that 77% had follow-up periods of at least 5 years [34]. In addition, deaths were confirmed via family interviews or healthcare referrals rather than death registries, potentially underestimating mortality. Study design differences may contribute to these inconsistent findings.

NEAT score was not significantly associated with mortality, cardiovascular events or cancer. Several considerations should be taken into account when interpreting these null findings, particularly those for mortality. The mortality analysis included only 32 deaths among 1,463 participants and was therefore likely underpowered to detect a modest association with habitual non-exercise PA. Two additional factors may have attenuated an underlying association. First, self-reported NEAT primarily captures low-intensity daily movement and is susceptible to measurement error. Such error may reduce the ability to distinguish differences in NEAT between participants and bias the estimated association toward the null. Second, the prognostic contribution of NEAT may have been less apparent in this population with diabetes, in whom mortality risk is strongly influenced by the substantial burden of established metabolic and CV comorbidities [35].

Exploratory analyses of additional body composition measures showed that higher SMM, SMI and phase angle were significantly associated with lower all-cause mortality, whereas FM and FFM were not. These findings are consistent with previous reports that low SMM has been associated with all-cause mortality in individuals with T2D [36,37] and that phase angle, a bioelectrical marker of cell-membrane integrity and nutritional status, has been reported as a prognostic marker of survival [38–40]. However, these associations were based on only 10 deaths and should therefore be considered hypothesis-generating. Larger cohorts with longer follow-up are needed to determine whether muscle-mass indices and phase angle independently predict mortality in individuals with diabetes.

To our knowledge, few studies have examined associations between VAT and hard clinical endpoints specifically in individuals with diabetes. Franssens et al. [41] reported no significant associations of intra-abdominal fat with cardiovascular events or mortality in T2D. In contrast, the present study identified a significant association between VAT and cancer incidence. The present study is also noteworthy for evaluating the associations of NEAT score with all-cause mortality, CV events and cancer incidence, although no significant associations were observed. NEAT encompasses the continuous, low-intensity PA accumulated throughout daily life and remains difficult to measure objectively, even with advanced technologies [42].

Several limitations should be acknowledged. First, measurements were unavailable for some participants. NEAT score, HGS and VAT measurements were available for 1,504, 1,472, and 1,106 participants, respectively, and approximately 900 participants had complete data for all three measures. The limited availability of these measurements reduced the precision of the analyses. VAT assessment required an additional fee and time, and each participant decided whether to undergo the assessment. Participants who chose to undergo VAT measurement were younger and had a shorter duration of diabetes than those who did not. Consequently, the VAT cohort represented a comparatively healthier subgroup of the overall study population. This difference may limit the generalisability of the VAT findings. The smaller cohort size and lower-risk profile may also have contributed to the small number of deaths in the VAT analysis. Second, although Cox proportional hazards models were adjusted for several relevant covariates, residual confounding by unmeasured factors, including dietary intake, medication use, comorbidities and socioeconomic status, cannot be excluded. These factors may influence both body composition and daily PA. Third, the study was conducted within a single healthcare organisation, which may limit the generalisability of the findings to other populations and clinical settings. Finally, cancer incidence was analysed as a composite outcome, and information on cancer subtype was unavailable. Therefore, the observed association between VAT and cancer incidence cannot be attributed to any specific cancer type. Because the relationship between visceral adiposity and cancer risk may differ by tumour site, future studies should investigate site-specific cancer incidence in individuals with diabetes.

## 5. Conclusion

In individuals with diabetes, higher HGS was independently associated with a lower risk of CV events, whereas higher VAT was associated with an increased risk of cancer incidence. NEAT score was not significantly associated with mortality, CV events or cancer incidence. These findings suggest that assessments of muscle strength and body composition may provide clinically relevant information beyond conventional anthropometric measures in individuals with diabetes. Further prospective studies are warranted to confirm these findings and determine their clinical utility in risk stratification and routine diabetes care.

## Declaration of competing interest

The authors declare that they have no competing interests that could have appeared to influence this research.

## Data Availability

All data produced in the present study are available upon reasonable request to the authors

## Acknowledgements

The authors thank the director and medical staff of Hamasaki Clinic for their support in conducting this study.

AI-assisted tools were used to support programming and English language editing. All statistical analyses, model specifications, interpretations, and conclusions were independently verified by the corresponding author.

## Notes

### Competing Interest Statement

The authors have declared no competing interest.

### Author Declarations

The study protocol received approval from the Japan Medical Association Ethical Review Board (reference number: JMA-IIA00340) and adhered to the principles of the Declaration of Helsinki.

### Summary of Updates

This revised version includes several methodological and reporting improvements.

