## Supplementary material for "Association of daily physical activity, handgrip strength and visceral adiposity with mortality, cardiovascular events and cancers in Japanese individuals with diabetes": Suppleentary Materials

### **Supplementary materials**

**NEAT questionnaire**

3 points are scored when participants answer "A lot".

2 points are scored when participants answer "Sometimes".

1 point is scored when participants answer "Hardly ever" or "No".

In question No.4,

3 points are scored when participants answer "Faster than most people".

2 points are scored when participants answer "About the same as other people".

1 point is scored when participants answer "I get passed by other people".

*Primarily walking physical exertion*

1. Do you walk on the way to work?

2. Do you use a train or a bus (not including taxis or private cars)?

3. Do you walk a lot while working?

4. Do you walk fast or slow?

5. Do you use stairs?

6. Do you go shopping for food or other daily necessities?

7. Do you walk to eat out (including lunch, and not including travel by car)?

8. Do you take the garbage out?

9. Do you go to concerts, to the theatre or karaoke?

10. Do you play with young children outside?

11. Do you ever go for a walk (including walking the dog)?

*Non-primarily walking physical exertion*

12. Do you ever do any light cleaning (such as picking up trash)?

13. Do you ever do any relatively hard cleaning (such as using a vacuum cleaner, mopping a floor, or dusting)?

14. Do you ever clean any large objects (such as windows, ventilation fans, or cars)?

15. Do you ever prepare meals (cooking or serving)?

16. During meals, do you often get up to fill rice bowls, or get things?

17. Do you clear the table?

18. Do you wash the dishes?

19. Do you wipe the dishes and put them away?

20. Do you do the washing (carrying and putting out to dry)?

21. Do you bring the washing in (and fold it up)?

22. Do you wash the bed sheets and covers?

23. Do you ever put the bedding outside in the sun?

24. Do you do the ironing?

25. Do you clean the bath and the toilet?

26. Do you clean the garden and around the house?

27. Do you ever do any weeding or gardening?

28. Do you water any plants?

29. Do you look after or feed any pets?

30. Do you have to look after any young children (such as cooking meals, dressing, or playing together inside)?

31. Do you ever pick them up (such as giving them a hug or a piggy-back ride)?

32. Do you have to look after anyone old or sick?

33. How often do you take a bath (or shower)?

34. Do you do any sewing or any other handcraft?

35. Do you play an instrument?

36. Do you ride a bicycle?

**Supplementary Table 1a.** Univariate linear regression analyses of non-exercise activity thermogenesis score and each parameter.

| **Parameter** | **Coefficient (B)** | **95% CI** | **p-value** |
| --- | --- | --- | --- |
| Age (years) | 0.026 | −0.016 to 0.068 | 0.23 |
| BMI (kg/m^2^) | −0.175 | −0.301 to −0.049 | 0.007 |
| WC (cm) | −0.109 | −0.163 to −0.055 | <0.001 |
| Systolic blood pressure (mmHg) | −0.002 | −0.033 to 0.029 | 0.92 |
| Diastolic blood pressure (mmHg) | −0.089 | −0.139 to −0.040 | <0.001 |
| Plasma glucose (mg/dL) | −0.004 | −0.012 to 0.003 | 0.25 |
| HbA1c (%) | 0.087 | −0.214 to 0.387 | 0.57 |
| TC(mg/dL) | 0.020 | 0.005 to 0.034 | 0.007 |
| TG (mg/dL) | −0.006 | −0.011 to −0.001 | 0.011 |
| HDL-C (mg/dL) | 0.124 | 0.087 to 0.161 | <0.001 |
| LDL-C (mg/dL) | 0.021 | 0.004 to 0.038 | 0.018 |
| C-peptide (ng/mL) | −0.124 | −0.830 to 0.582 | 0.73 |
| HGS (kg) | −0.241 | −0.297 to −0.185 | <0.001 |
| FM (kg) | 0.001 | −0.085 to 0.088 | 0.98 |
| Body fat percentage (%) | 0.270 | 0.195 to 0.345 | <0.001 |
| FFM (kg) | −0.367 | −0.428 to −0.307 | <0.001 |
| Lean body mass percentage (%) | −0.198 | −0.265 to −0.130 | <0.001 |
| SMM (kg) | −0.619 | −0.724 to −0.515 | <0.001 |
| SMI | −1.983 | −2.354 to −1.613 | <0.001 |
| VAT (L) | −2.006 | −2.581 to −1.431 | <0.001 |
| Phase angle | −2.927 | −3.815 to −2.039 | <0.001 |

NEAT, non-exercise activity thermogenesis; CI, confidence interval; BMI, body mass index; WC, waist circumference; HbA1c, haemoglobin A1c; TC, total cholesterol; TG, triglycerides; HDL-C, high-density lipoprotein cholesterol; LDL-C, low-density lipoprotein cholesterol; HGS, handgrip strength; FM, fat mass; FFM, fat-free mass; SMM, skeletal muscle mass; SMI, skeletal muscle mass index; VAT, visceral adipose tissue volume.

**Supplementary Table 1b.** Univariate linear regression analyses of handgrip strength and each parameter.

| **Parameter** | **Coefficient (B)** | **95% CI** | **p-value** |
| --- | --- | --- | --- |
| Age (years) | −0.339 | −0.376 to −0.302 | <0.001 |
| BMI (kg/m^2^) | 0.442 | 0.324 to 0.560 | <0.001 |
| WC (cm) | 0.091 | 0.040 to 0.142 | <0.001 |
| Systolic blood pressure (mmHg) | −0.017 | −0.046 to 0.013 | 0.26 |
| Diastolic blood pressure (mmHg) | 0.245 | 0.199 to 0.291 | <0.001 |
| Plasma glucose (mg/dL) | 0.005 | −0.003 to 0.012 | 0.22 |
| HbA1c (%) | 0.235 | −0.052 to 0.522 | 0.11 |
| TC(mg/dL) | 0.000 | −0.014 to 0.013 | 0.95 |
| TG (mg/dL) | 0.013 | 0.008 to 0.017 | <0.001 |
| HDL-C (mg/dL) | −0.121 | −0.156 to −0.086 | <0.001 |
| LDL-C (mg/dL | −0.006 | −0.022 to 0.010 | 0.44 |
| C-peptide (ng/mL) | 0.056 | −0.621 to 0.732 | 0.87 |
| NEAT | −0.223 | −0.275 to −0.171 | <0.001 |
| FM (kg) | −0.103 | −0.181 to −0.024 | 0.011 |
| Body fat percentage (%) | −0.574 | −0.633 to −0.514 | <0.001 |
| FFM (kg) | 0.698 | 0.659 to 0.738 | <0.001 |
| Lean body mass percentage (%) | 0.470 | 0.413 to 0.527 | <0.001 |
| SMM (kg) | 1.253 | 1.188 to 1.318 | <0.001 |
| SMI | 4.053 | 3.800 to 4.305 | <0.001 |
| VAT (L) | 1.978 | 1.440 to 2.516 | <0.001 |
| Phase angle | 8.847 | 8.206 to 9.489 | <0.001 |

HGS, handgrip strength; CI, confidence interval; BMI, body mass index; WC, waist circumference; HbA1c, haemoglobin A1c; TC, total cholesterol; TG, triglycerides; HDL-C, high-density lipoprotein cholesterol; LDL-C, low-density lipoprotein cholesterol; NEAT, non-exercise activity thermogenesis; FM, fat mass; FFM, fat-free mass; SMM, skeletal muscle mass; SMI, skeletal muscle mass index; VAT, visceral adipose tissue volume.

**Supplementary Table 1c.** Univariate linear regression analyses of visceral fat tissue volume and each parameter.

| **Parameter** | **Coefficient (B)** | **95% CI** | **p-value** |
| --- | --- | --- | --- |
| Age (years) | −0.006 | −0.011 to −0.001 | 0.021 |
| BMI (kg/m^2^) | 0.173 | 0.163 to 0.184 | <0.001 |
| WC (cm) | 0.083 | 0.079 to 0.086 | <0.001 |
| Systolic blood pressure (mmHg) | 0.008 | 0.005 to 0.012 | <0.001 |
| Diastolic blood pressure (mmHg) | 0.023 | 0.017 to 0.029 | <0.001 |
| Plasma glucose (mg/dL) | 0.002 | 0.001 to 0.003 | 0.002 |
| HbA1c (%) | 0.031 | −0.006 to 0.068 | 0.10 |
| TC(mg/dL) | −0.002 | −0.004 to −0.000 | 0.018 |
| TG (mg/dL) | 0.002 | 0.001 to 0.002 | <0.001 |
| HDL-C (mg/dL) | −0.024 | −0.028 to −0.019 | <0.001 |
| LDL-C (mg/dL | −0.002 | −0.004 to 0.000 | 0.098 |
| C-peptide (ng/mL) | 0.246 | 0.170 to 0.322 | <0.001 |
| HGS (kg) | 0.027 | 0.020 to 0.034 | <0.001 |
| NEAT | −0.025 | −0.032 to −0.018 | <0.001 |
| FM (kg) | 0.095 | 0.089 to 0.101 | <0.001 |
| Body fat percentage (%) | 0.033 | 0.026 to 0.041 | <0.001 |
| FFM (kg) | 0.055 | 0.050 to 0.061 | <0.001 |
| Lean body mass percentage (%) | −0.025 | −0.032 to −0.019 | <0.001 |
| SMM (kg) | 0.096 | 0.087 to 0.106 | <0.001 |
| SMI | 0.357 | 0.325 to 0.389 | <0.001 |
| Phase angle | 0.376 | 0.289 to 0.464 | <0.001 |

VAT, visceral adipose tissue volume; CI, confidence interval; BMI, body mass index; WC, waist circumference; HbA1c, haemoglobin A1c; TC, total cholesterol; TG, triglycerides; HDL-C, high-density lipoprotein cholesterol; LDL-C, low-density lipoprotein cholesterol; NEAT, non-exercise activity thermogenesis; HGS, handgrip strength; FM, fat mass; FFM, fat-free mass; SMM, skeletal muscle mass; SMI, skeletal muscle mass index.

**Supplementary Table 2.** Associations of additional body-composition indices with all-cause mortality, cardiovascular events, and cancer.

| **Body-composition index** | **Outcome** | **n** | **Events** | **HR (95% CI)** | **p-value** |
| --- | --- | --- | --- | --- | --- |
| FM | All-cause mortality | 1,102 | 10 | 0.928 (0.841–1.024) | 0.14 |
| FM | CV events | 852 | 45 | 1.030 (0.990–1.072) | 0.15 |
| FM | Cancer | 938 | 39 | 1.041 (0.995–1.089) | 0.085 |
| FFM | All-cause mortality | 1,102 | 10 | 0.936 (0.830–1.054) | 0.27 |
| FFM | CV events | 852 | 45 | 1.017 (0.971–1.064) | 0.48 |
| FFM | Cancer | 938 | 39 | 1.039 (0.987–1.095) | 0.14 |
| SMM | All-cause mortality | 1,102 | 10 | 0.803 (0.654–0.985) | 0.035 |
| SMM | CV events | 852 | 45 | 1.023 (0.942–1.111) | 0.59 |
| SMM | Cancer | 938 | 39 | 1.065 (0.971–1.168) | 0.18 |
| SMI | All-cause mortality | 1,102 | 10 | 0.449 (0.266–0.760) | 0.003 |
| SMI | CV events | 852 | 45 | 1.178 (0.908–1.528) | 0.22 |
| SMI | Cancer | 938 | 39 | 1.280 (0.965–1.698) | 0.086 |
| Phase angle | All-cause mortality | 1,102 | 10 | 0.171 (0.066–0.441) | <0.001 |
| Phase angle | CV events | 852 | 45 | 0.871 (0.509–1.490) | 0.61 |
| Phase angle | Cancer | 938 | 39 | 1.064 (0.598–1.891) | 0.83 |

HR, hazard ratio; CI, confidence interval; FM, fat mass; FFM, fat-free mass; SMM, skeletal muscle mass; SMI, skeletal muscle mass index; CV, cardiovascular. Each index was analysed as a continuous variable in a separate Cox proportional hazards model. Mortality models were adjusted for age and sex only, because the small number of deaths precluded the fully adjusted model (see Table 3 footnote and Methods); cardiovascular event and cancer models were adjusted for age, sex, BMI, smoking and drinking habits, regular exercise, systolic blood pressure, HDL-C, LDL-C, HbA1c and diabetes duration. Fine-Gray subdistribution hazard models treating death as a competing event yielded concordant results for cardiovascular events and cancer (all p > 0.05).

**Supplementary Table 3.** Differences in clinical parameters between participants divided by the quartile of visceral fat tissue volume.

|  | **Q1** | **Q2** | **Q3** | **Q4** |  | **p-value** | **p-value (post-hoc test)** |
| --- | --- | --- | --- | --- | --- | --- | --- |
| Age (years) | 58.6 (14.1) | 63.2 (11.6) | 62.2 (11.8) | 58.8 (14.1) |  | <0.001 | <0.001^a^  0.007 ^b^  <0.001 ^e^  0.008 ^f^ |
| Sex |  |  |  |  |  | <0.05 | <0.05 ^b,c,d,e,f^ |
| Men | 88 (36.8%) | 128 (45.9%) | 204 (55.9%) | 230 (83.0%) |  |  |  |
| Women | 151 (63.2%) | 151 (54.1%) | 161 (44.1%) | 47 (17.0%) |  |  |  |
| Smoking habit |  |  |  |  |  | <0.05 | <0.05 ^b,c,d,e^ |
| Current or past | 53 (24.9%) | 70 (26.3%) | 118 (40.7%) | 133 (51.6%) |  |  |  |
| Never | 160 (75.1%) | 196 (73.7%) | 172 (59.3%) | 125 (48.4%) |  |  |  |
| Drinking habit |  |  |  |  |  | <0.05 | <0.05 ^e^ |
| Current or past | 110 (52.1%) | 118 (45.0%) | 160 (55.4%) | 164 (63.6%) |  |  |  |
| Never | 101 (47.9%) | 144 (55.0%) | 129 (44.6%) | 94 (36.4%) |  |  |  |
| Regular exercise habit |  |  |  |  |  | <0.05 | <0.05 ^c,e^ |
| Yes | 95 (51.6%) | 101 (45.9%) | 106 (42.4%) | 78 (33.5%) |  |  |  |
| No | 89 (48.4%) | 119 (54.1%) | 144 (57.6%) | 155 (66.5%) |  |  |  |
| History of CV disease |  |  |  |  |  | NS | NS |
| Yes | 21 (9.6%) | 46 (17.2%) | 40 (13.6%) | 38 (14.4%) |  |  |  |
| No | 198 (90.4%) | 222 (82.8%) | 254 (86.4%) | 225 (85.6%) |  |  |  |
| Height (cm) | 159.2 (8.9) | 159.7 (8.7) | 162.9 (8.7) | 166.3 (8.1) |  | <0.001 | <0.001 ^b,c,d,e,f^ |
| Weight (kg) | 54.4 (10.4) | 60.8 (8.9) | 68.2 (9.8) | 81.2 (14.8) |  | <0.001 | <0.001 ^a,b,c,d,e,f^ |
| BMI (kg/m^2^) | 21.4 (3.2) | 23.9 (3.0) | 25.7 (3.4) | 29.3 (4.8) |  | <0.001 | <0.001 ^a,b,c,d,e,f^ |
| Waist circumference (cm) | 77.8 (7.5) | 86.2 (6.1) | 92.3 (6.3) | 102.1 (9.8) |  | <0.001 | <0.001 ^a,b,c,d,e,f^ |
| HGS (kg) | 25.7 (9.5) | 26.5 (9.3) | 29.7 (9.6) | 32.4 (10.2) |  | <0.001 | <0.001 ^b,c,e^  0.002 ^d^  0.013 ^f^ |
| NEAT score | 66.5 (9.9) | 66.2 (10.1) | 62.1 (10.2) | 59.4 (10.3) |  | <0.001 | <0.001 ^b,c,d,e^  0.002 ^f^ |
| Systolic blood pressure (mmHg) | 122.4 (18.3) | 128.1 (19.1) | 129.1 (16.6) | 129.9 (16.9) |  | <0.001 | 0.002 ^a^  <0.001 ^b,c^ |
| Diastolic blood pressure (mmHg) | 68.8 (10.2) | 71.2 (11.3) | 73.5 (11.0) | 75.4 (11.5) |  | <0.001 | <0.001 ^b,c,e^ |
| ABI | 1.16 (0.080) | 1.15 (0.087) | 1.15 (0.086) | 1.15 (0.086) |  | 0.56 |  |
| TBI | 0.82 (0.16) | 0.79 (0.15) | 0.81 (0.14) | 0.82 (0.14) |  | 0.27 |  |
| baPWV (cm/s) | 1658 (381) | 1676 (344) | 1692 (328) | 1618 (373) |  | 0.22 |  |
| CVRR | 3.0 (1.6) | 3.0 (1.4) | 3.3 (2.3) | 3.3 (2.0) |  | 0.48 |  |
| TC (mg/dL) | 206.9 (40.8) | 213.0 (40.9) | 205.4 (41.7) | 200.6 (37.9) |  | 0.007 | 0.003 ^e^ |
| TG (mg/dL) | 121.9 (126.2) | 136.8 (86.1) | 168.2 (149.3) | 188.0 (129.6) |  | <0.001 | <0.001 ^b,c,e^  0.018 ^d^ |
| HDL-C (mg/dL) | 62.1 (16.7) | 56.7 (14.7) | 52.4 (13.6) | 47.6 (12.5) |  | <0.001 | <0.001 ^a,b,c,f^  0.003 ^e^ |
| LDL-C (mg/dL) | 121.2 (36.3) | 128.1 (35.5) | 119.5 (33.2) | 116.7 (34.9) |  | 0.003 | 0.033 ^d^  0.002 ^e^ |
| Plasma glucose (mg/dL) | 156.9 (67.8) | 157.9 (66.8) | 166.2 (65.8) | 171.4 (68.7) |  | 0.047 |  |
| HbA1c (%) | 7.8 (2.2) | 8.0 (2.0) | 7.8 (1.8) | 8.0 (1.8) |  | 0.41 |  |
| C-peptide (ng/mL) | 1.7 (1.5) | 2.0 (1.3) | 2.4 (1.4) | 2.8 (2.0) |  | <0.001 | <0.001 ^c^  0.006 ^e^ |
| UACR (mg/gCr) | 48.6 (185.4) | 69.1 (184.9) | 44.4 (100.7) | 47.3 (104.7) |  | 0.36 |  |
| FM (kg) | 15.6 (6.1) | 19.8 (5.9) | 22.9 (6.1) | 28.7 (8.4) |  | <0.001 | <0.001 ^a,b,c,d,e,f^ |
| Body fat percentage (%) | 29.0 (10.5) | 33.2 (9.8) | 33.9 (8.4) | 35.2 (6.8) |  | <0.001 | <0.001 ^a,b,c^  0.038 ^e^ |
| FFM (kg) | 38.2 (9.4) | 40.0 (8.9) | 45.2 (9.0) | 52.4 (9.9) |  | <0.001 | <0.001 ^b,c,d,e,f^ |
| Lean body mass percentage (%) | 70.4 (11.4) | 65.7 (11.3) | 66.0 (8.8) | 64.4 (8.1) |  | <0.001 | <0.001 ^a,b,c^ |
| SMM (kg) | 15.9 (5.4) | 17.2 (5.2) | 20.3 (5.2) | 24.3 (5.7) |  | <0.001 | 0.031 ^a^  <0.001 ^b,c,d,e,f^ |
| SMI (kg/m^2^) | 6.1 (1.6) | 6.6 (1.5) | 7.5 (1.4) | 8.7 (1.5) |  | <0.001 | 0.002 ^a^  <0.001 ^b,c,d,e,f^ |
| VAT (L) | 1.9 (0.3) | 2.6 (0.2) | 3.2 (0.2) | 4.6 (1.1) |  | <0.001 | <0.001 ^a,b,c,d,e,f^ |
| Phase angle | 4.5 (0.7) | 4.6 (0.7) | 4.8 (0.7) | 5.0 (0.8) |  | <0.001 | <0.001 ^b,c,d,e^  0.022 ^f^ |

To detect significant differences between the groups divided by the quartile of VAT, a one-way analysis of variance (ANOVA) test was performed. Tukey’s post-hoc test was conducted when the ANOVA identified a statistically significant difference. Study participants were divided into Q1 (0.4 L ≤ VAT < 2.3L), Q2 (2.3 L ≤ VAT < 2.9 L), Q3 (2.9 L ≤ VAT < 3.7 L) and Q4 (VAT ≥ 3.7 L). One-way ANOVA revealed that age, weight, BMI, WC, HGS, systolic blood pressure, diastolic blood pressure, blood glucose levels, TC levels, TG levels, HDL-C levels, LDL-C levels, C-peptide levels, NEAT score, FM, body fat percentage, FFM, lean body mass percentage, SM, SMI and phase angle significantly differed between the groups divided by the quartile of VAT. Additionally, the sex ratio, smoking and drinking habits and the presence or absence of regular exercise habits were significantly different between the groups divided by the quartiles of VAT. Data are represented as mean (SD) except for number of participants, sex, cardiovascular disease history, smoking, drinking and regular exercise habits. CV, cardiovascular; BMI, body mass index; HGS, handgrip strength; NEAT, non-exercise activity thermogenesis; ABI, ankle–brachial index; TBI, toe–brachial index, baPWV, brachial–ankle pulse wave velocity; CVRR, the coefficient of variation of R–R intervals; TC, total cholesterol; TG, triglycerides; HDL-C, high-density lipoprotein cholesterol; LDL-C, low-density lipoprotein cholesterol; HbA1c, hemoglobin A1c; UACR, urinary albumin creatinine ratio; FM, fat mass; FFM, fat-free mass; SMM skeletal muscle mass, SMI, skeletal muscle mass index; VAT, visceral fat tissue volume; NS, not significant.

^a^ significant differences between Q1 and Q2. ^b^ significant differences between Q1 and Q3. ^c^ significant differences between Q1 and Q3. ^d^ significant differences between Q2 and Q3. ^e^ significant differences between Q2 and Q4. ^f^ significant differences between Q3 and Q4.
